# Consensus guideline for the management of patients with appendiceal tumors Part 1: Appendiceal tumors without peritoneal involvement

**DOI:** 10.1101/2024.04.09.24305468

**Authors:** PSM Appendiceal Tumor Writing Group, PSM Consortium Group, Kiran K Turaga

## Abstract

**Background:** Appendiceal tumors comprise a heterogeneous group of tumors which may be localized or disseminate throughout the peritoneum. Limited high quality clinical data exists and many practices have been extrapolated from colorectal cancer without validation in appendiceal cohorts. Many controversies exist regarding their treatment, and practices vary widely between centers and care settings. A national consensus update of best management practices for appendiceal malignancies was performed to better standardize care.

**Methods:** The 2018 Chicago consensus guideline was updated via modified Delphi consensus, performed over two rounds using nationally circulated surveys. Supporting evidence was evaluated using rapid systematic reviews. Key systemic therapy concepts were summarized by content experts.

**Results:** Most supporting literature consists of observational studies, but increasingly high-quality studies are becoming available to drive management. Two consensus-based pathways were generated for localized appendiceal tumors, one for epithelial mucinous neoplasms and another for appendiceal adenocarcinoma. Of 138 participants responding in the first round, 133 (96%) engaged in the second round. Over 90% consensus was achieved for all pathway blocks. Key points include minimizing intervention invasiveness where permitted by pathologic classification and margin status, and determining what margin and pathologic findings are indications for consideration of cytoreduction with or without intraperitoneal chemotherapy. Surveillance and systemic therapy recommendations are also presented.

**Conclusion:** With growing but still primarily observational evidence currently dictating care, these consensus recommendations provide expert guidance in the treatment of appendiceal tumors without peritoneal involvement.

## BACKGROUND

Appendiceal tumors comprise a diverse group of pathologies of the vermiform appendix. Their incidence has been markedly increasing, doubling in the years 2004-2017 alone; recent estimates report 0.97 cases per 100,000 individuals.^1,2,3^ Although still a rare disease, it is critical for general surgeons to be familiar with appendix tumors because of the substantially higher incidence of 1-3% in those undergoing appendectomy, and for primary and emergency care generalists to avoid missed diagnoses.^4–7^ Complicated appendicitis, including perforation or abscess, is associated with greater risk of a neoplastic diagnosis, with rates ranging from 5-29%. ^5,8–12^

Mucinous neoplasms represent over half of appendix tumors; the rest are predominantly epithelial (65-70%) followed by neuroendocrine (∼20%).^13^ The most common epithelial malignancies are mucinous adenocarcinoma (35-40%), followed by colonic/intestinal type (7-27%), goblet cell (about 20%), and signet ring adenocarcinoma (estimates usually under 10%). ^1,14,15^ Approximately 40-50% of appendix tumors present with distant disease at diagnosis, usually peritoneal. ^3,14,16–19^

Prognosis varies widely across disease histology and stage. Low grade neuroendocrine tumors, not addressed by this guideline, have the best prognosis; of non-metastatic epithelial tumors, the most recent studies report five-year overall survival of 63-75% for well- and moderately-differentiated mucinous disease, and 60-70% for non-mucinous.^1,3,14,17,18,20–23^ Data on non-metastatic, higher-grade tumors is scant as they often present at more advanced stages.

Given the rarity of appendiceal tumors, prospective studies are challenging and randomized studies are nearly non-existent, so data to guide their management are low-quality and there are no well-established standards of care.^24,25^ To fill this need, the multidisciplinary Chicago Consensus Working Group was formed in 2018 to generate consensus recommendations for peritoneal malignancies including appendix tumors.^26^ Herein, these recommendations are updated by expert consensus for the clinical management of patients with localized appendiceal mucinous neoplasms and localized appendiceal adenocarcinoma, supported with recent evidence synthesized through rapid systematic reviews.

### Conceptual Overview and Changes from the 2018 Chicago Consensus^26^

Peritoneal disease has been removed from the pathology-defined localized pathways and reorganized as a unified treatment pathway, which will be addressed in a separate document. All pathways feature a more comprehensive, multidisciplinary initial evaluation recommendation. Pathways have been streamlined to emphasize preferred treatment options. Surveillance recommendations have been unified across pathways. Finally, systemic chemotherapy tables have been developed to describe prevailing trends in systemic treatment.

## METHODS

The methods for the 2023 consensus update of the 2018 Chicago Consensus Guidelines have been described in detail in a separate manuscript.^27^ Major components are presented below.

### Consensus Group Structure

The Appendiceal Tumor Working Group included fifteen multidisciplinary experts. Two steering committee core members coordinated the effort and prepared all revisions (FM, EG). Sixteen trainees (medical students, residents, and fellows) conducted the rapid reviews.

### Modified Delphi Process

The original Chicago Consensus guidelines were reviewed by the Appendiceal Tumor Working Group and Consortium leadership to align with evidence published since the last consensus. Recommendations were revised using two rounds of modified Delphi consensus across the Consortium by soliciting degrees of agreement with each recommendation on a five-point Likert scale via Qualtrics survey. A threshold of 75% was set for inclusion of a guideline, with revision required below and considered above 90% consensus to improve agreement.

### Rapid Review of the Literature

Rapid systematic reviews were performed of Pubmed indexed literature in Medline in three key areas, developed in conjunction with a medical librarian specialist. The search period ranged up to August 2023. The search strategies and study protocol were registered with the international prospective register of systematic reviews (PROSPERO) and therefore will not be replicated here. The search strategy may be reviewed in the supplement (Supplemental Table 1).

The following key review question is addressed in this document:

1. In patients with moderate to poorly differentiated appendiceal adenocarcinoma undergoing cytoreductive surgery, which systemic therapy sequences and regimens are associated with superior survival and safety outcomes (total neoadjuvant, perioperative, adjuvant alone)? (PROSPERO CRD42023463216)

The other two key questions will be discussed in the part 2 appendiceal tumor guidelines with the accompanying peritoneal disease pathway.

Reviews were conducted and data extracted according to the published review methodology with minimal changes. Further criteria emerging from screening may be reviewed in the supplement (Supplemental Table 2). As no randomized trials were eligible for inclusion, quality analysis utilized the Newcastle Ottawa framework, which allots up to nine stars for methodologic quality, with six or higher considered good-quality.^28,29^ Abstract and full text screening was performed in duplicate, and extraction and quality analysis was performed individually with secondary verification.

The systemic chemotherapy table presented herein was drafted collaboratively by the Appendiceal Tumor Working Group, with directed guidance from medical oncologist contributors. It was then circulated for feedback from the consortium group alongside the Delphi round 2 consensus survey.

## RESULTS

### Pathways

Of 138 experts who voted on the clinical pathways for appendiceal mucinous neoplasms (AMN) and appendiceal adenocarcinoma in the first round, 133 (96%) participated in the second round. The group comprised 92 (67%) surgical oncologists, 20 (16%) medical oncologists, 12 (12%) pathologists, and 5 (5%) experts from other disciplines. This pathway was divided into eleven main blocks. After two Delphi rounds, the blocks are summarized below with supporting literature incorporated where appropriate.

### Rapid review

A total of 1179 abstracts were screened; 247 were included for full-text review and a total of 34 were selected for inclusion in the review, reporting outcomes specific to patients with peritoneal metastases of moderate- and poorly-differentiated appendiceal origin undergoing cytoreductive surgery and systemic chemotherapy. Exclusions are quantified in the PRISMA flow diagram (Figure 1) and further described in Supplemental Table 2. Outcomes were overall and disease-free survival, and adverse events. Seventeen studies reported on preoperative or neoadjuvant chemotherapy (^30–46^), 6 reported on postoperative or adjuvant chemotherapy (^47–51^), 9 reported on both (^52–60^), and 2 reported on other or unspecified regimens (^61,62^). These studies are summarized in Table 2, addressed qualitatively in Principles of Systemic Therapy, and applied in blocks 3 and 6 of the Appendiceal Adenocarcinoma pathway. Quality assessment can be found in Supplemental Table 3.

**Figure 1.**
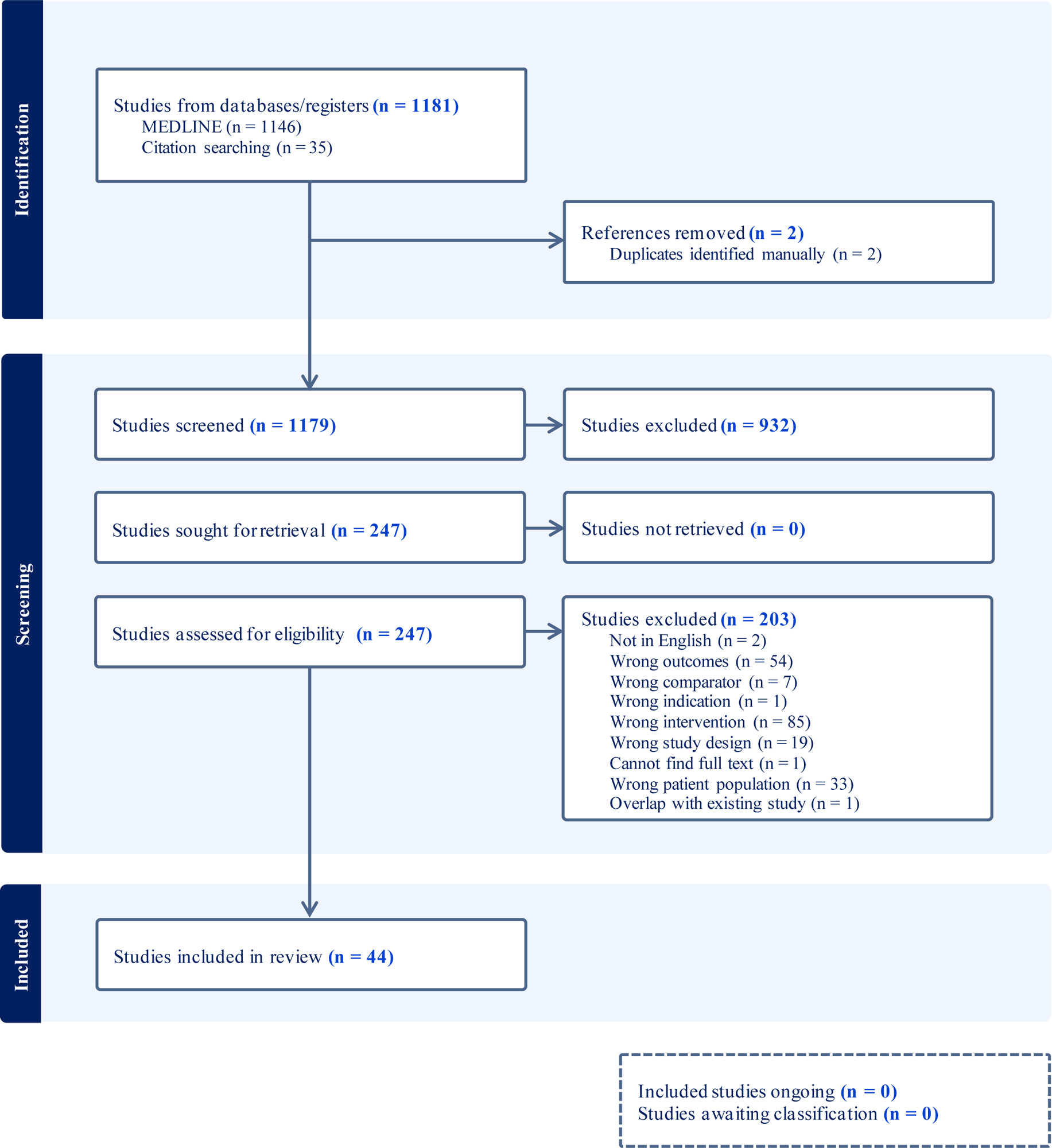
PRISMA Diagram.

## PRINCIPLES OF SYSTEMIC THERAPY

One of the key current issues in appendix tumors is the role, regimen, and timing of systemic chemotherapy, which is currently influenced by a combination of the few small single prospective or retrospective studies in appendix tumors, and larger trials in colorectal cancer, despite increasing evidence that appendix cancer has a distinct biology from colorectal cancer.^63–67^ In addition to the lack of high-quality evidence, low-grade lesions are also likely resistant to systemic chemotherapy, confounding the results of prior studies and limiting the applicability of their conclusions.^68^

### Cytotoxic chemotherapy for localized appendix tumors

Localized appendix tumors may include WHO grade 1 primaries, either low-grade appendiceal mucinous lesions or well-differentiated adenocarcinoma. High-grade appendiceal mucinous neoplasms most likely should be treated similarly. At present, available studies indicate no benefit from the use of 5-FU based chemotherapy in this population, so it is not recommended.^24,53,69–71^

When the primary lesion is an adenocarcinoma with high-risk features without peritoneal involvement, up-front resection with consideration of adjuvant chemotherapy is preferred by expert consensus. High risk features have largely been extrapolated from colorectal cancer literature without validation in appendiceal cohorts, including T4 tumor size, invasion of adjacent structures, inadequate lymph node yield, and tumor perforation. High-risk features validated in appendiceal malignancy include lymph node involvement, signet ring cells, and less differentiated or non-mucinous histology.^52,54,71–77^

The rationale for this is partly mechanistic, as tumors with poor biology are anticipated to have higher likelihood of distant spread. Limited observational evidence has shown benefit associated with the use of adjuvant chemotherapy in node-positive, high-grade, and/or non-mucinous disease.^48,53,54,59,60,71–73,78^ Others studies have shown minimal benefit or even detriment after adjuvant therapy; it is unclear how much of this variation reflects selection bias, as the patients who are most likely to undergo adjuvant therapy are those who are well enough to do so.^47,49,51,52,56^ To our knowledge, no studies have comprehensively evaluated the role of modern neoadjuvant therapy for resectable appendix tumors without peritoneal involvement; at present expert consensus opinion is not in favor of neoadjuvant therapy in that setting, as it would delay definitive resection.

### Cytotoxic chemotherapy for tumors with peritoneal disease

Pathologic grade typically dictates the role of chemotherapy in appendix tumors with peritoneal spread. If both the primary tumor and associated peritoneal lesions are low-grade, cytotoxic therapies are usually not indicated. When resectable, definitive resection should be pursued. When not resectable, palliative debulking may be considered. Cytotoxic systemic therapies may be a part of clinical trials or in care pathways focused on symptom control, but no evidence currently supports their use for improved disease control or survival. If the peritoneal disease is low-grade but the primary is found to have high-risk features, as above, adjuvant chemotherapy should be considered as for any high-grade primary.

For appendiceal tumors with high-grade peritoneal disease histology, the grade of the primary does not affect management; even where there is substantial discordance, such as a LAMN or well-differentiated adenocarcinoma (which would be vanishingly rare), the peritoneal pathology guides management. Some studies suggest some degree of disease response with systemic chemotherapy, with disease stability or improvement on imaging in 20-75% of patients, and some patients with unresectable disease becoming eligible for cytoreduction.^79–82^ In one prospective trial, 50% of 34 patients receiving preoperative chemotherapy had disease stability or response on imaging, confirmed by intraoperative findings, and of those 17, 53% (9 of the 17) had lower tumor grade on pathology than in samples from prior chemotherapy.^32^ A subset of observational studies support modest disease control or response and increased survival after preoperative chemotherapy.^31–34,37,39,61^

However, this may not translate to cohort-wide overall-, recurrence-free, or progression-free survival, as a number of observational studies suggest a lack of benefit of preoperative chemotherapy in one or all of those domains.^31,33,34,36,41,42,44,47,50,52,54,56,57,83^ Overall and disease-free survival is still poor even with definitive cytoreduction; a large study of the US HIPEC collaborative estimates 23.2% five-year disease-free survival and 43.8% overall survival for high-grade appendiceal tumors with peritoneal involvement.^84^ Some studies show a survival benefit from postoperative therapy as well, but there is conflicting data regarding its role or benefit.^48,52,53^ This observation suggests that disease-specific underlying features which are poorly understood may be driving these treatment outcomes. Further research in this area may allow for directed management.

Weighing the existing evidence summarized above and in Table 2, which summarizes all studies included in the key question 1 rapid review, the expert consensus recommendation of the Peritoneal Surface Malignancy Consortium is to administer chemotherapy prior to attempting cytoreduction, or as definitive therapy if cytoreduction is not feasible, for high-grade peritoneal malignancy of appendiceal origin. When complete cytoreduction is predicted, systemic chemotherapy is useful for assessing disease biology and response, and when incomplete cytoreduction is predicted (high PCI or other anatomic factors), it is recommended as conversion therapy. If cytoreduction is incomplete or if the preoperative regimen was incomplete, postoperative chemotherapy should be considered. There is no clear consensus on regimen timing; when studied, perioperative regimens have been shown to potentially be more challenging for patients to complete than total preoperative, but may be worth considering, particularly when surgery must be expedited.^85^

### Cytotoxic chemotherapy regimens

Systemic chemotherapy for appendiceal malignancy commonly relies on the intravenous 5-fluorouracil backbone, or less commonly oral capecitabine, typically used in colorectal cancers. Regimens are typically either doublet, with oxaliplatin or irinotecan as the second agent, or triplet, with both; in patients unable to tolerate doublet or triplet chemotherapy, singlet may be employed.^35,37–39,46,48,53,57,73,81,86,87^ At this time only small retrospective studies have been done, so there is no clear evidence suggesting better outcomes with either regimen, but there is higher toxicity with triplet regimens, mandating careful patient selection.^38^ Most therapeutic regimens paired with definitive surgical management, whether pre-, peri-, or postoperative, are intended for a 3-6 month duration, but if definitive surgical management is not feasible, cytotoxic chemotherapy may be part of a long-term management strategy. Re-evaluation is generally performed every three months when intended to query disease biology or attempt conversion to resectable disease.^26,32,57^

### Appendix tumor genetics and targeted and molecular therapies

The role of targeted and molecular therapies is not well-defined in appendix tumors and is still largely extrapolated from colorectal and other GI cancers, but recent studies have explored genetic profiles of appendiceal tumors in the hopes of identifying effective targets. Four of the most common mutations in appendix cancer are KRAS (>70%), GNAS (50-70%), TP53 (up to 40%), and APC (up to 20%). The relative frequency of these mutations is distinct from that of colorectal cancer, particularly in the paucity of APC and TP53 mutations compared to 70-80% of colorectal cancers. High microsatellite-instability and MMR-deficiency are relatively uncommon in appendix cancer (6%) as well.^65–67,88,89^

As with most solid tumors, all patients with metastatic disease should receive next generation sequencing for molecular profiling with an accepted next generation sequencing panel to identify potential molecular targets. Retrospective data suggests that molecular information may also inform prognosis and/or predict therapy response, although targeted randomized studies in appendiceal cancer have not been performed.^64,65^ When possible, tissue should be sent for tumor molecular profiling; circulating (blood) profiling may not be as sensitive.^90^

Germline variants, including those associated with hereditary cancer syndromes, have been detected at frequencies approaching 10-12% in patients with appendiceal tumors, although these variants may be incidental to disease biology and the relevance to therapeutic management is unknown. ^89,90^ Testing for germline variants may be considered, taking into account the individual’s family history of cancer.

One molecularly-targeted treatment that may be applicable to metastatic appendiceal cancer is anti-VEGF agents, most commonly bevacizumab, which has been associated with improved outcomes in some observational studies.^87^ Anti-VEGF therapy may be considered in most settings in which systemic therapy is considered, with preference to those in which no resection or incomplete resection has taken place, although they should be avoided in patients assessed to be at risk of impending bowel obstruction or perforation, bleeding, or arterial thrombosis. Anti-EGFR agents have a more controversial role, as they have unclear survival benefit in appendix cancer and studies have raised concern for worse survival in patients with RAS mutations.^87,88^

Possible therapeutic options for less common mutations may be extrapolated from other cancers. The NCCN guidelines for appendix cancer at time of writing are presented alongside colorectal cancer recommendations and recommend similar use of targeted therapies for druggable targets in late, previously treated, and/or metastatic settings, such as treating BRAF V600E mutated tumors with combination anti-EGFR and anti-BRAF agents. ^91^ Deficient MMR and MSI-H lesions may be treated with anti-PD1 or combination anti-PD1 and anti-CTLA4 therapy.^91^ A recent trial investigated the effect of combination anti-PD1 (atezolizumab) and anti-VEGF (bevacizumab) therapy in 16 individuals with unresectable, predominantly low-grade mucinous appendiceal adenocarcinoma; disease control was achieved in 100% of individuals, with a PFS of 18 months compared to 3 months of disease control on 5-FU based regimens. This is a promising development for those with low-grade, unresectable disease.^92^

Genetic profiles of appendiceal tumors may also influence the effectiveness of cytotoxic regimens. Patients with GNAS-mutation predominant disease are much less likely to have a disease response to chemotherapy, while as many as 50% of patients with RAS-mutation predominant disease may respond.^65^ Additionally, some evidence may support the preferential use of irinotecan-containing regimens in RAS-wild type cancers.^64^

**Table 1.**
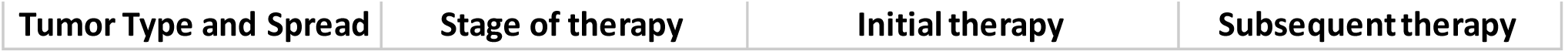
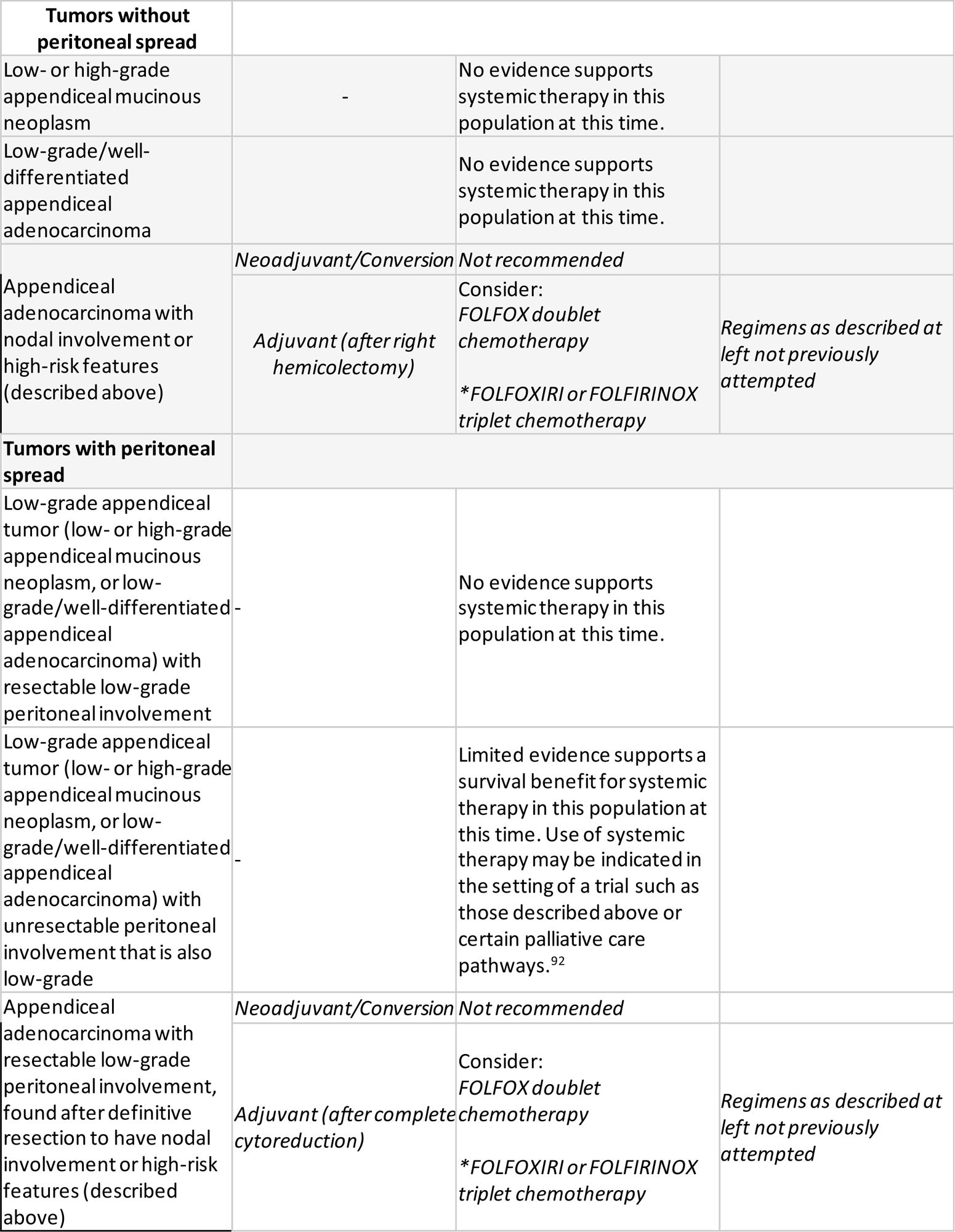
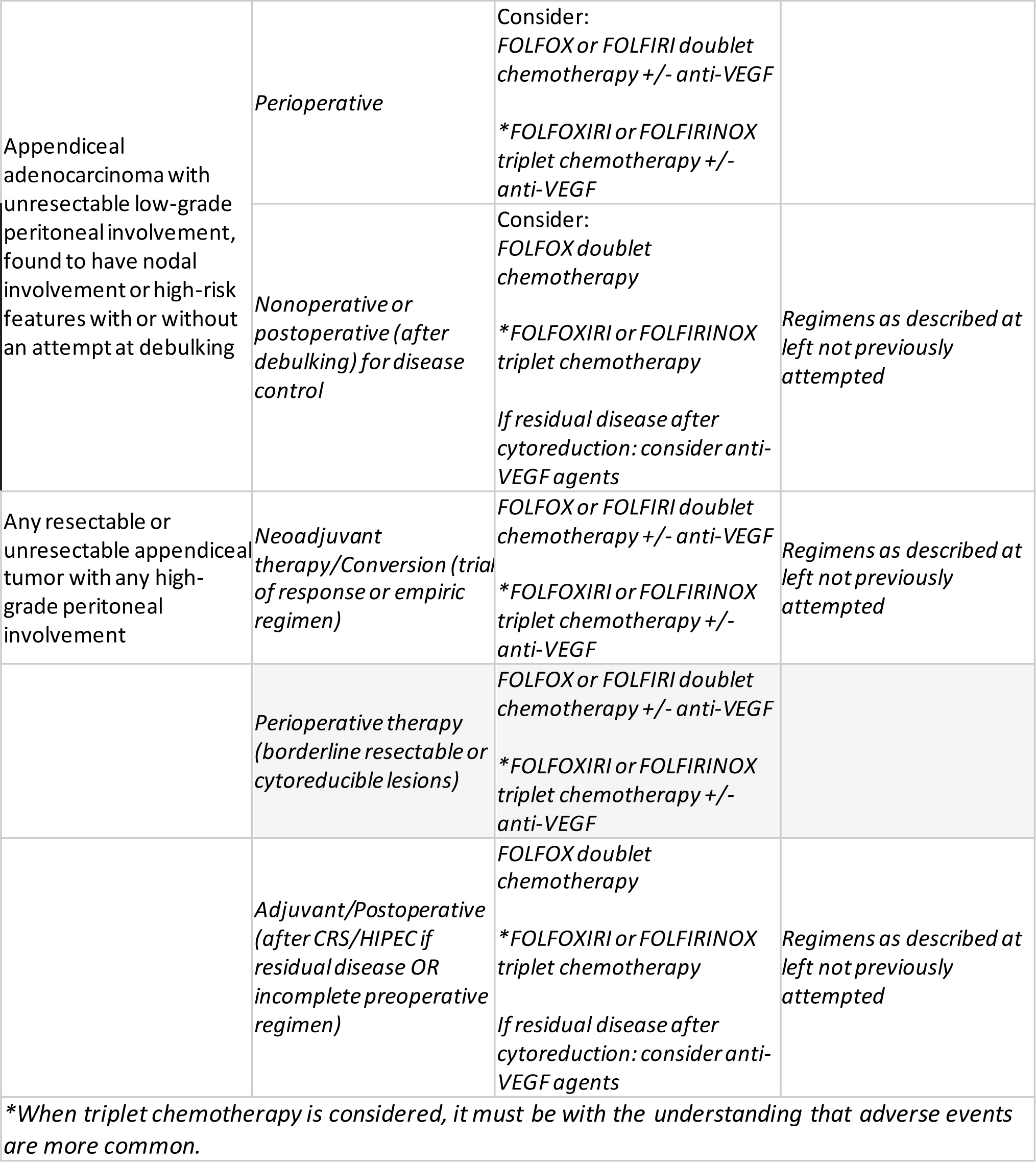
Systemic Chemotherapy for Appendiceal Tumors.

**Table 2.**
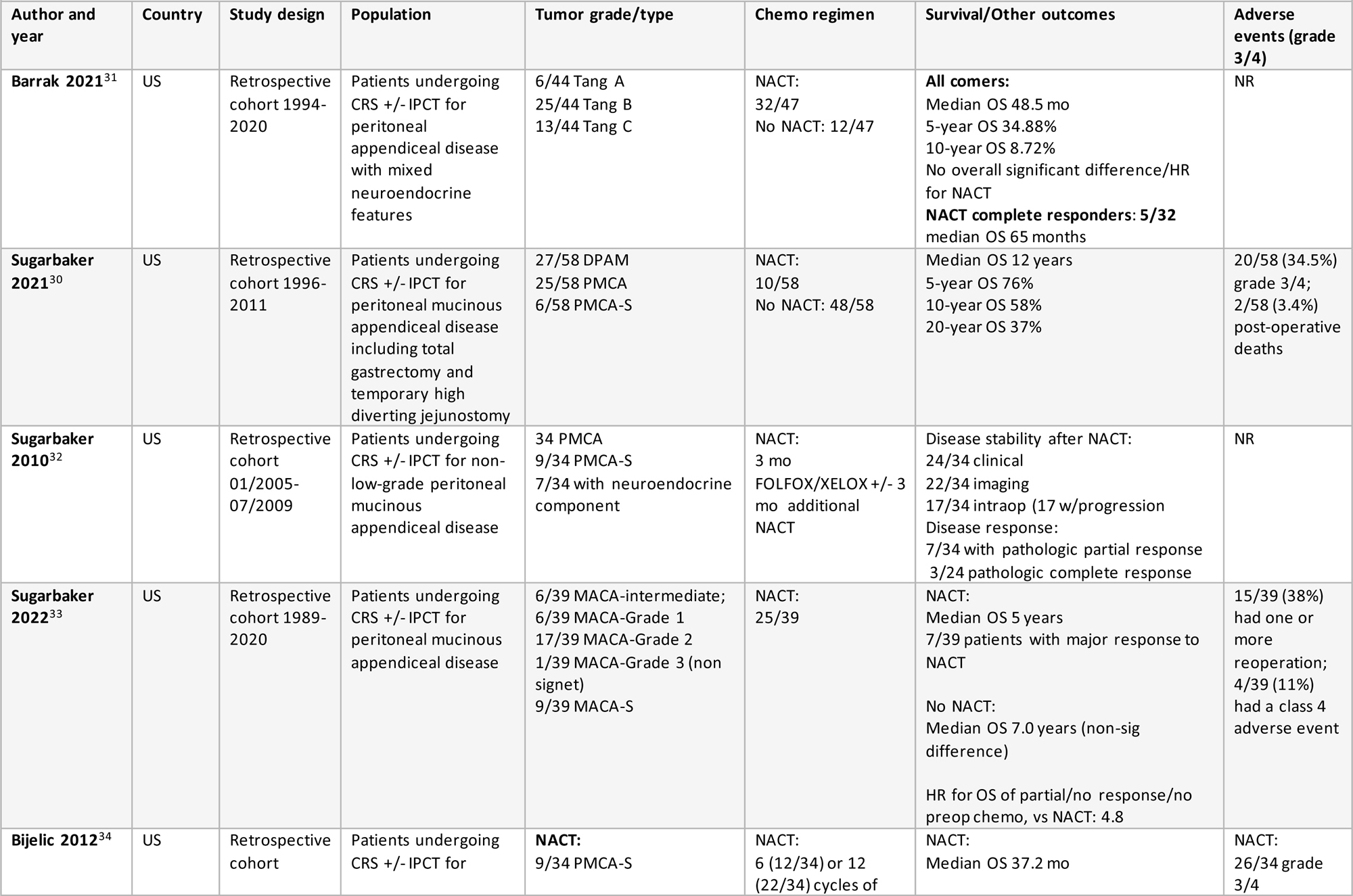
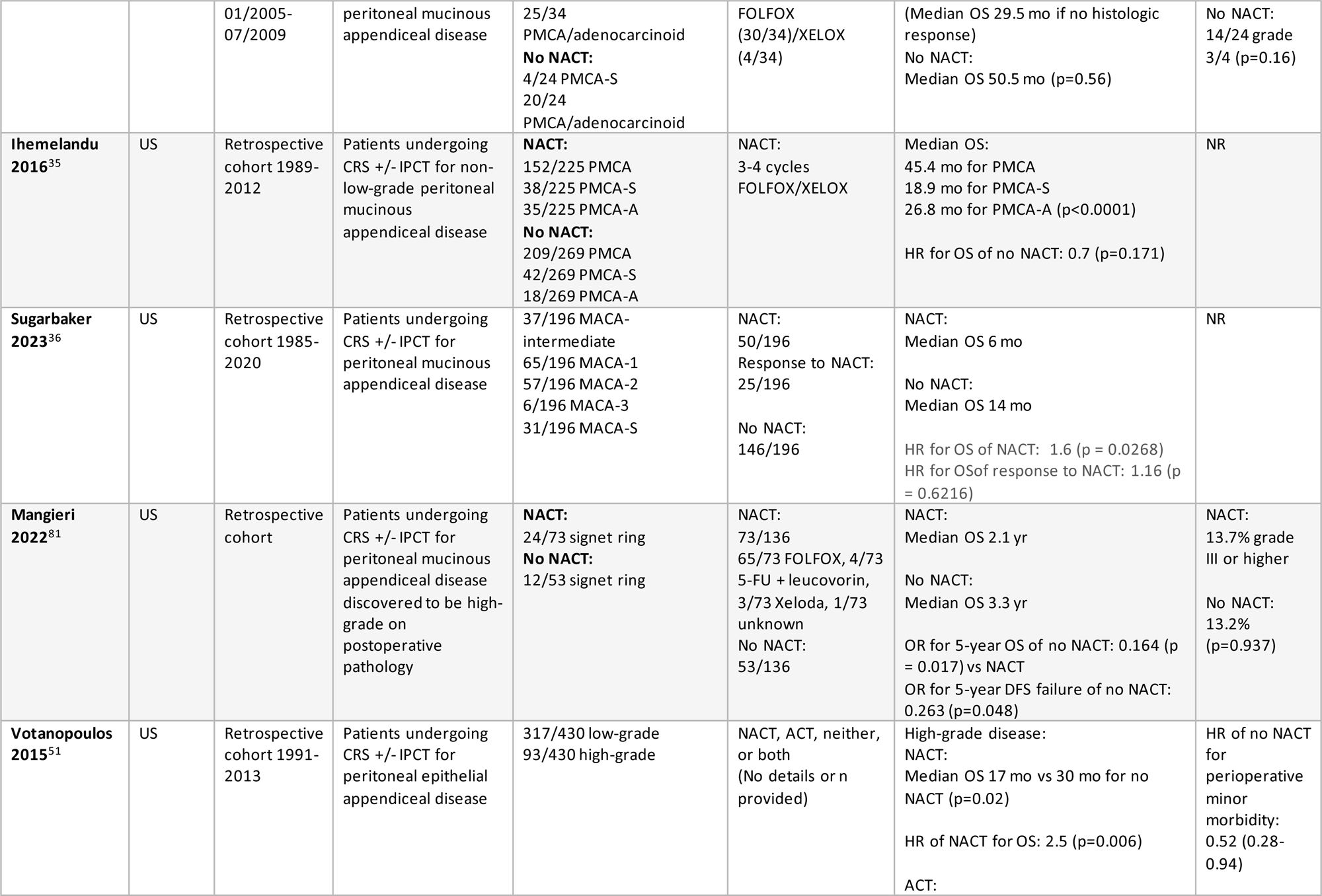
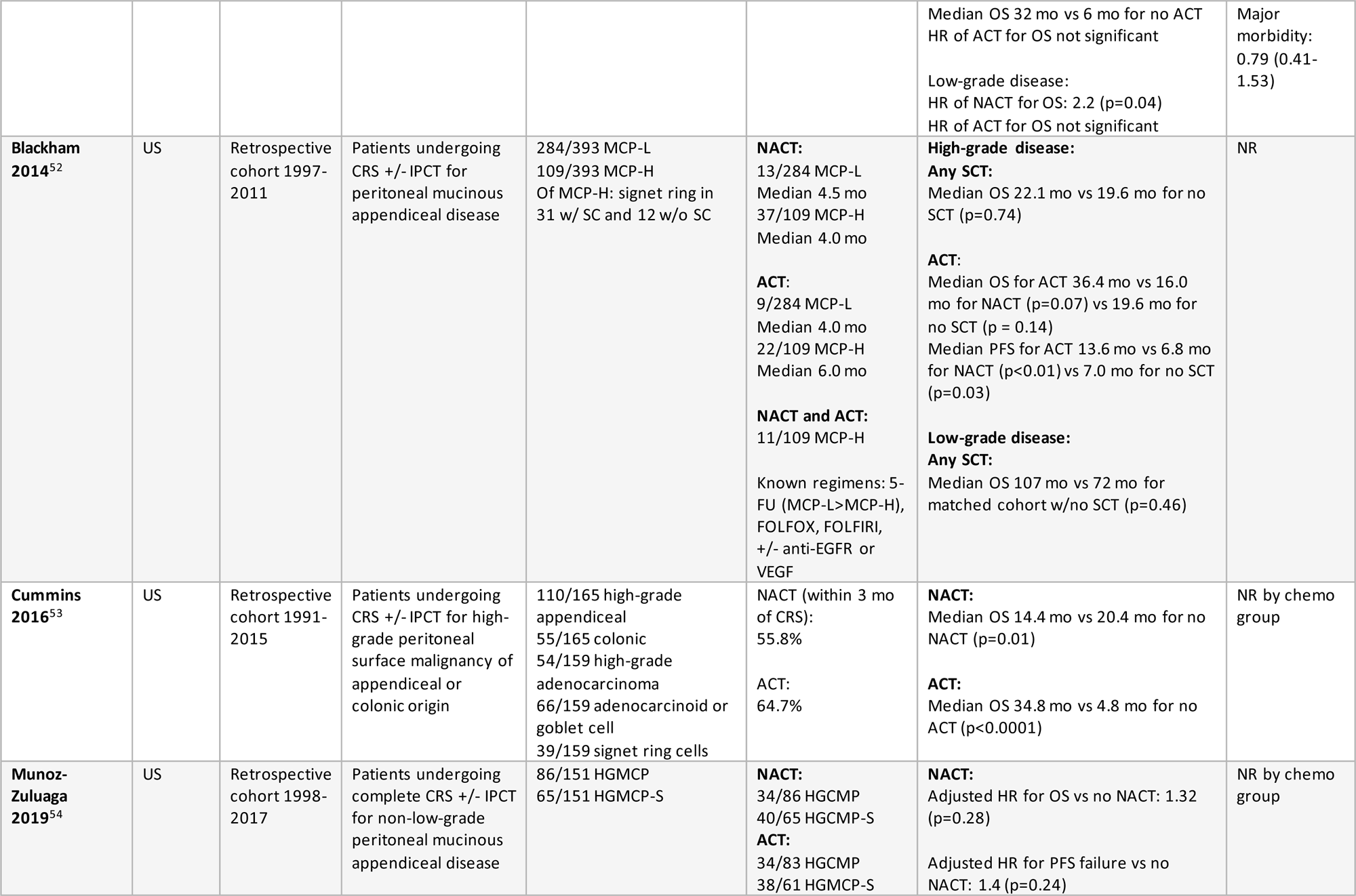
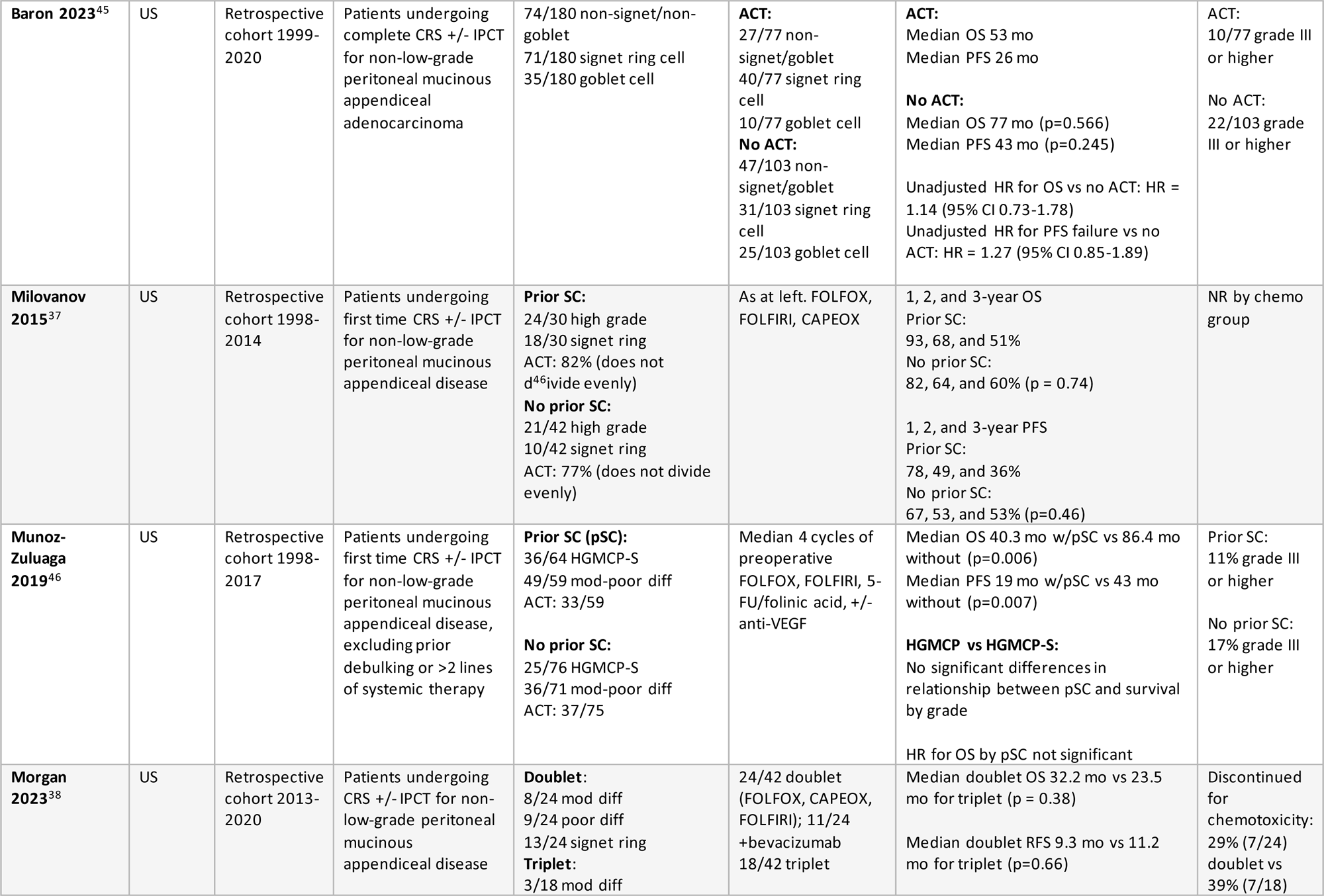
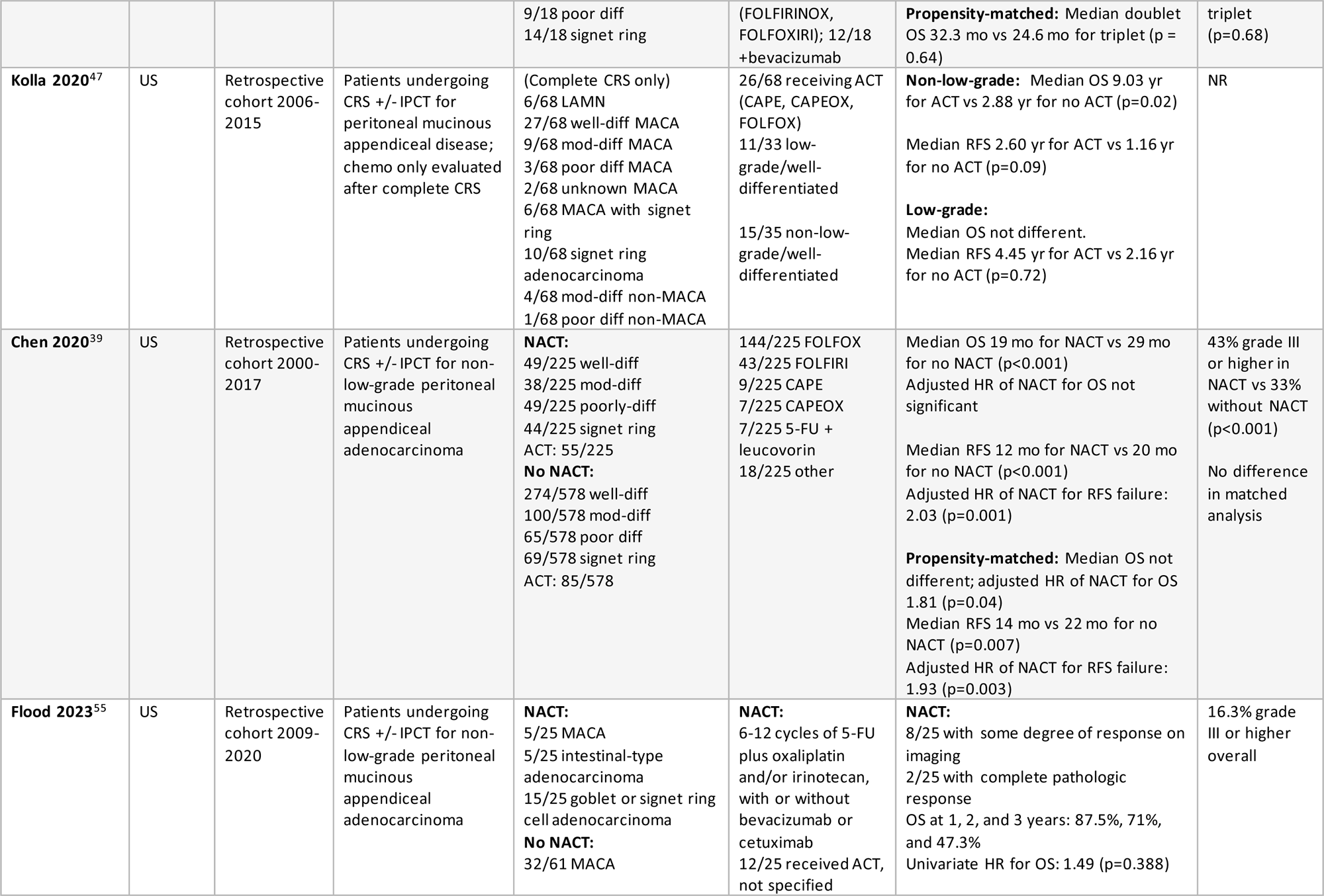
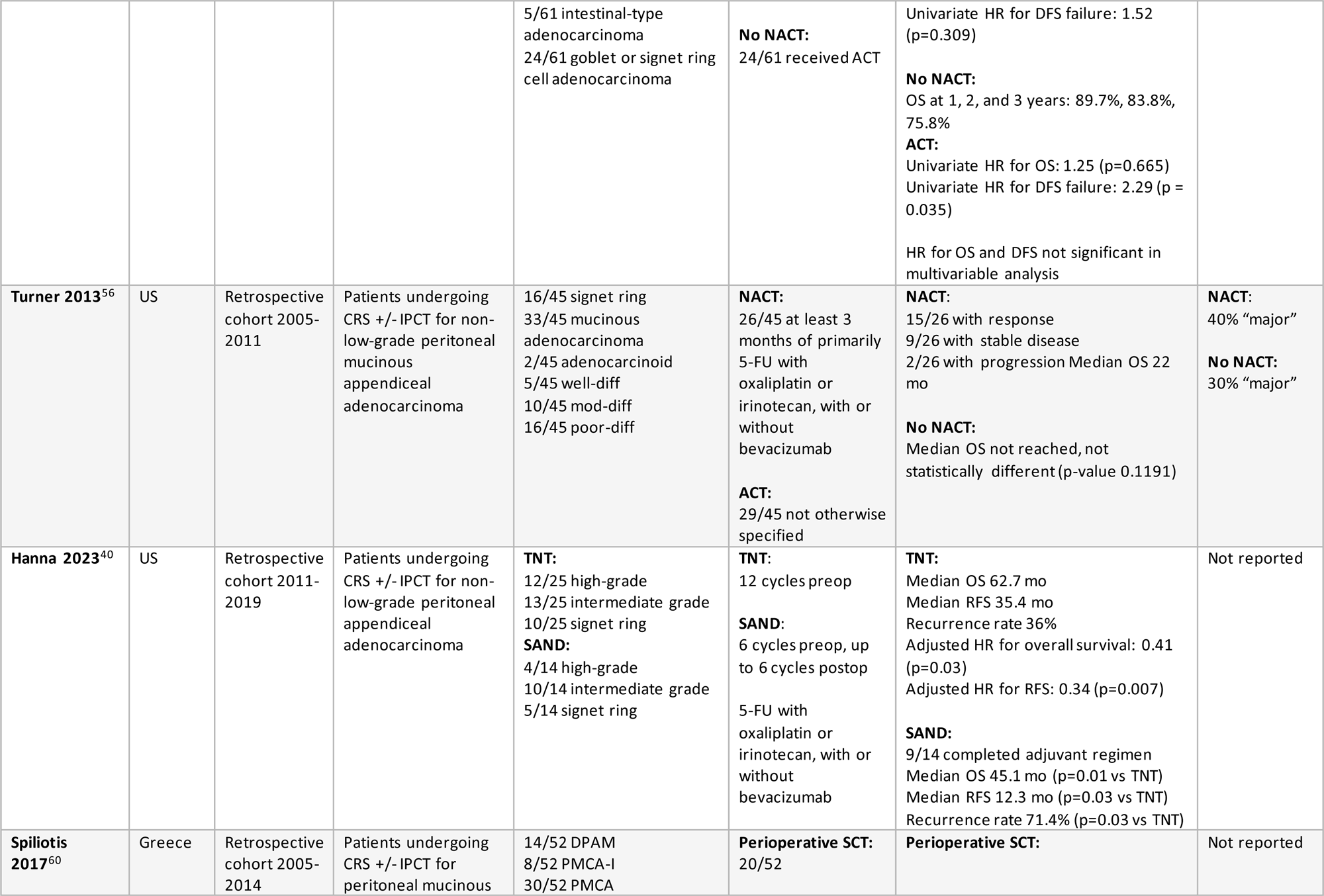
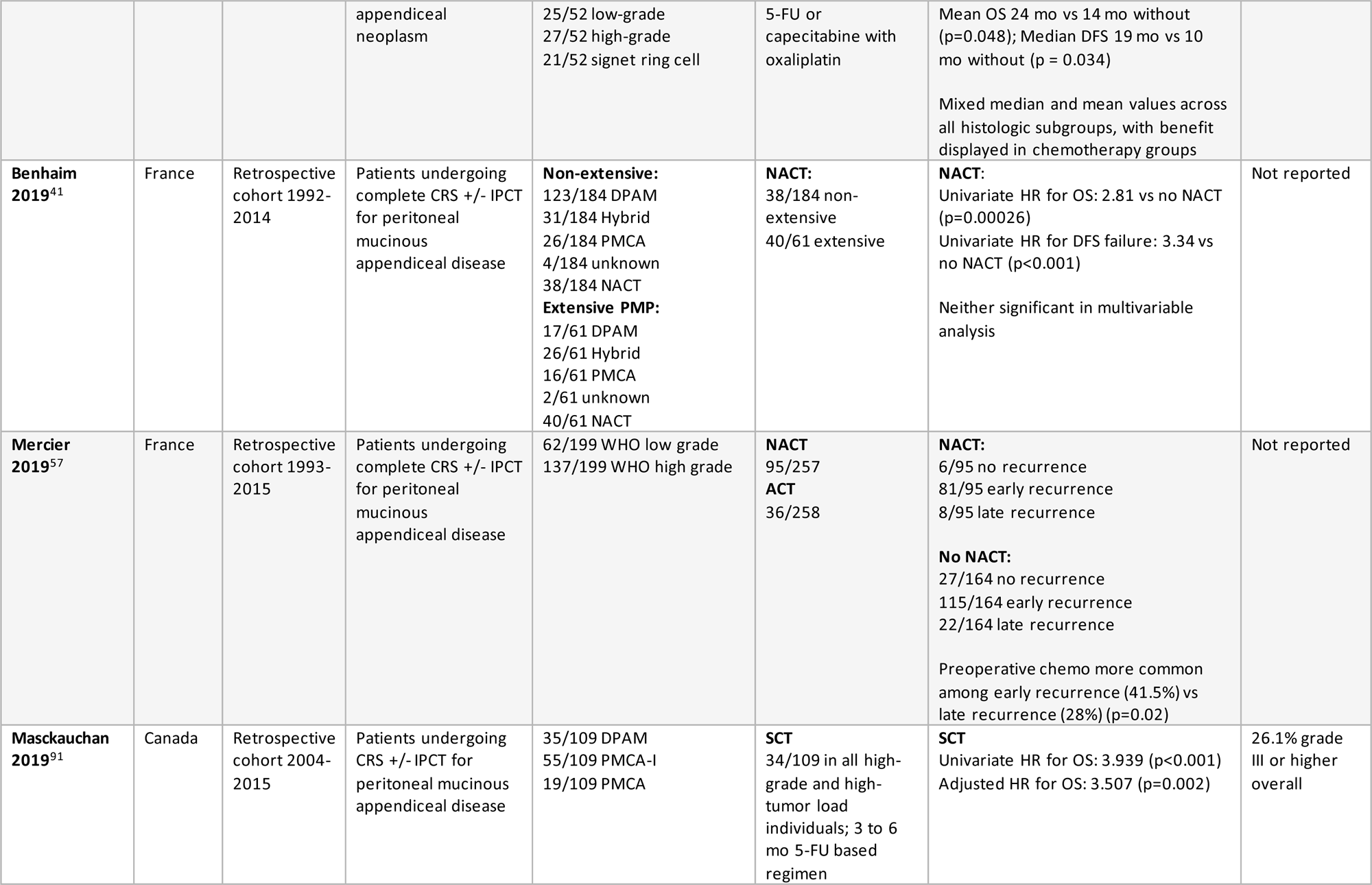
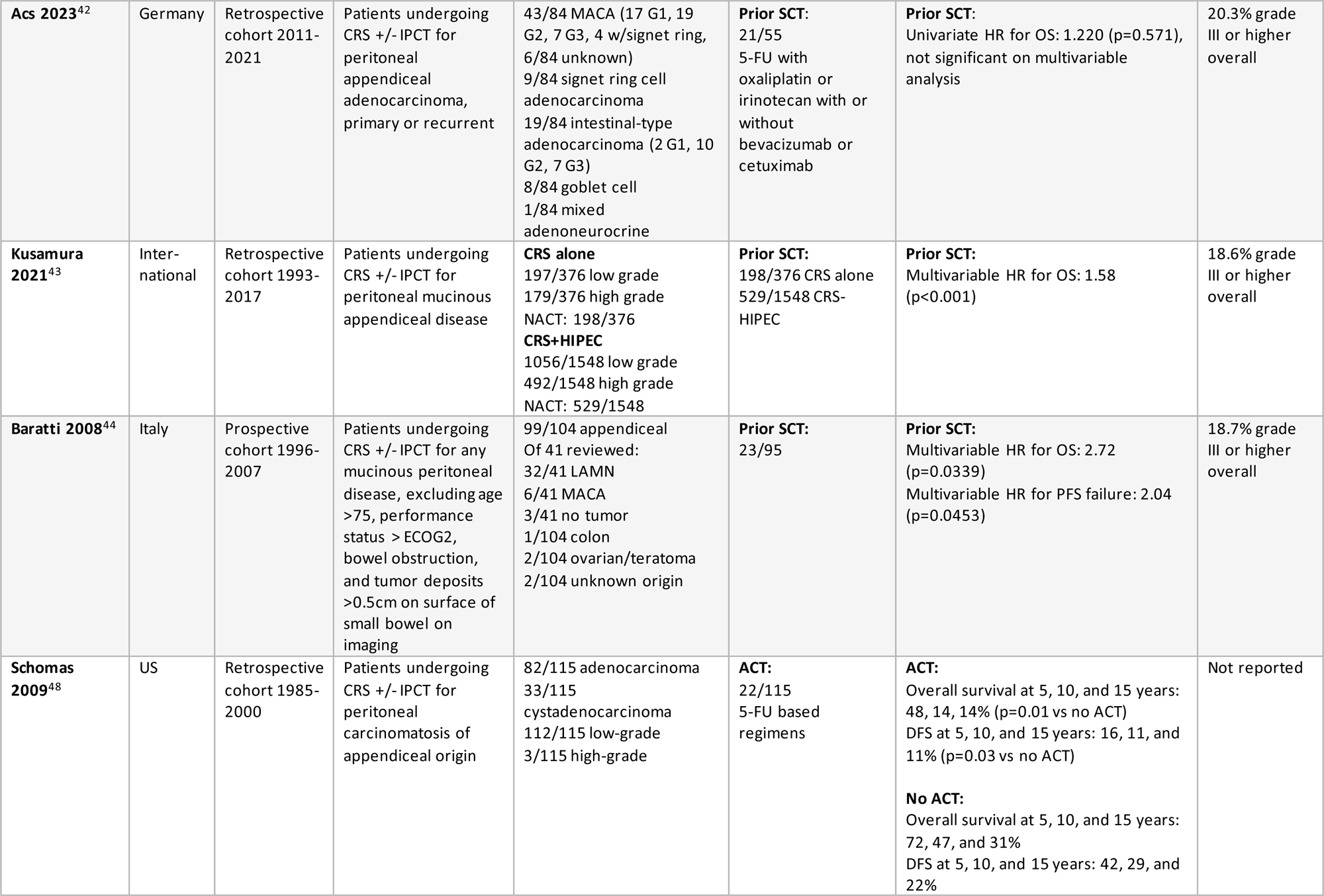
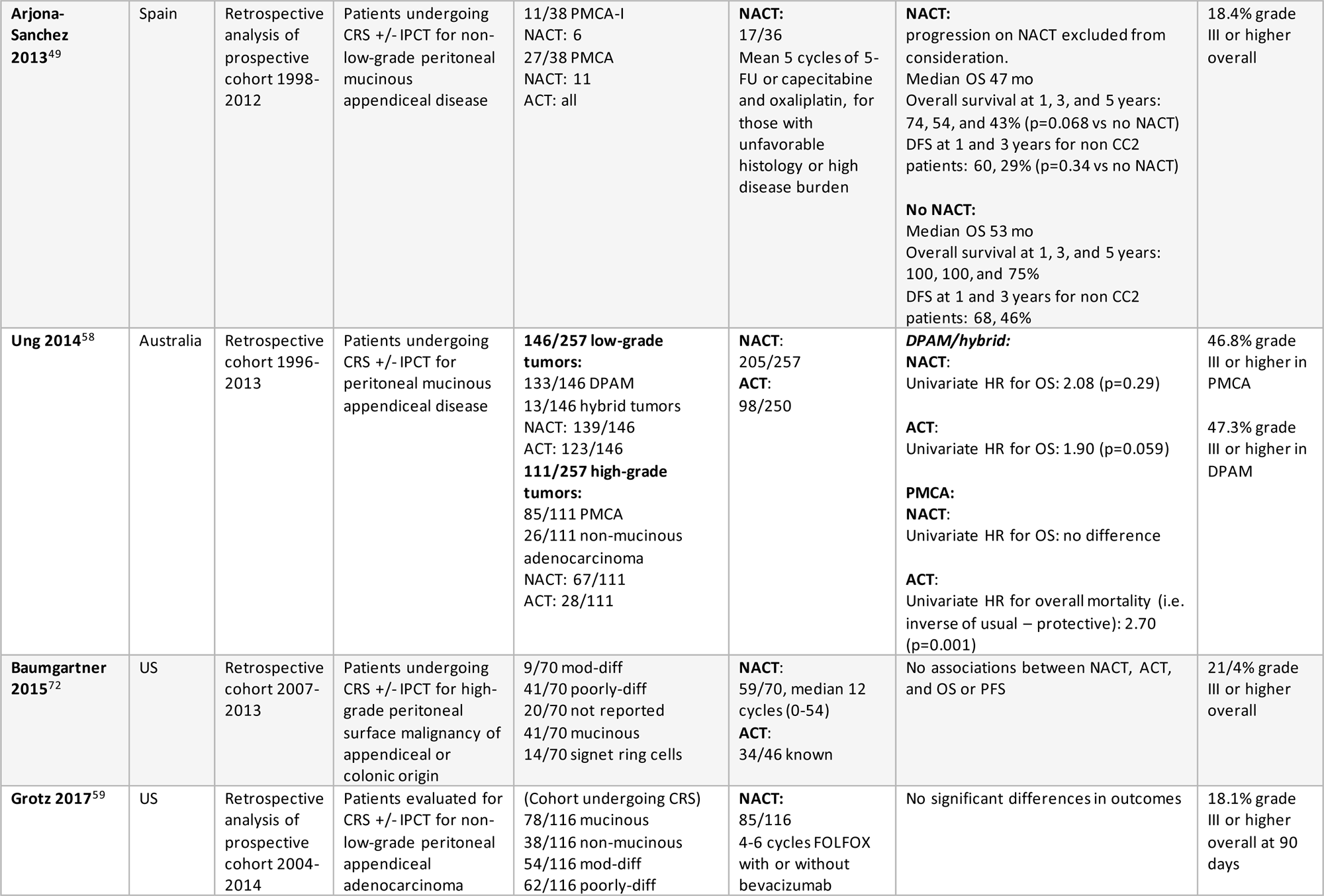
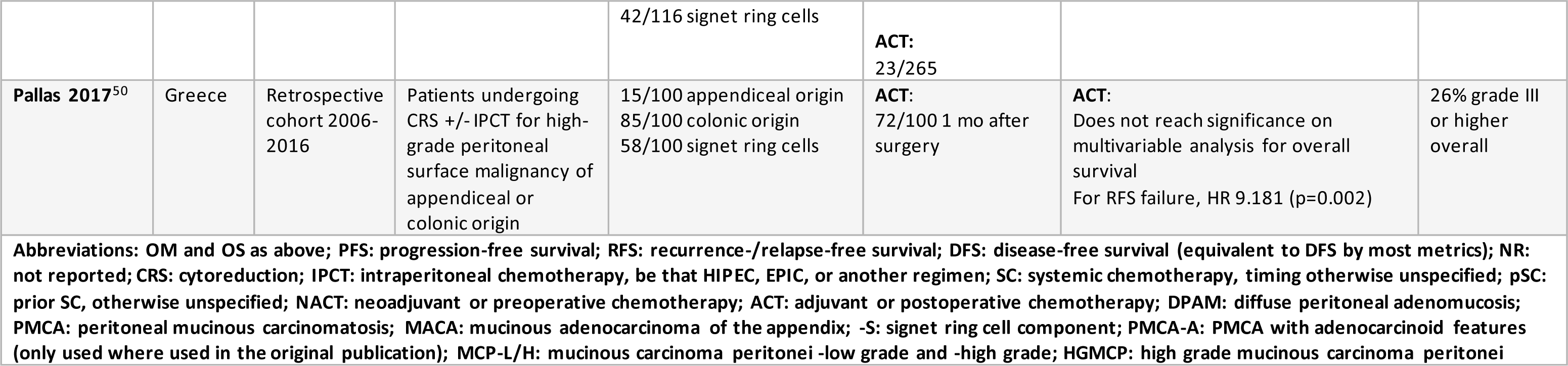
Key Question 1: Systemic chemotherapy regimens and timing relative to cytoreduction in peritoneal appendiceal malignancy.

### Regional chemotherapy regimens

Evidence suggests potential survival benefit from intraperitoneal chemotherapy with optimal cytoreduction for appendiceal neoplasms with peritoneal involvement.^35,42,43,69,86,93–96^ In general, the consensus recommendation is to consider intraperitoneal chemotherapy with optimal cytoreduction, but there are still variations in practice.

Mitomycin C (MMC) is the most widely used agent. ^35,69,75,93,96–110^ Oxaliplatin is also common, given its known activity against gastrointestinal malignancies.^42,94,95,99,111^ Oxaliplatin and MMC appear to have similar hematological outcomes, including in a randomized trial; MMC was more commonly associated with leukopenia and oxaliplatin with thrombocytopenia, with no difference in grade 3 and 4 adverse events.^99^ A few centers have studied regimens involving of irinotecan, cisplatin, and doxorubicin alone or in addition to MMC or oxaliplatin.^43,75,108,111–116^ Data is mixed regarding cisplatin-containing regimens but irinotecan trends toward more inferior outcomes. ^42,111^

In studies and centers performing pressurized intraperitoneal aerosol chemotherapy (PIPAC), oxaliplatin is most common, followed by cisplatin and doxorubicin.^117,118^ Currently, this consortium recommends PIPAC only in the setting of a clinical trial as early phase trials are still in progress. A small number of centers offer early postoperative intraperitoneal chemotherapy (EPIC); when implemented, 5-FU is typically used, with some initially promising data.^42,119,120^ The ICARUS and other trials are ongoing to further assess the role for EPIC in appendiceal cancer.^121^

**Table 3.** Regional Chemotherapy for Appendiceal Neoplasms.

| Regional Regimens | Currently in use |
| --- | --- |
| **HIPEC** | Mitomycin C<br>Oxaliplatin<br>Not recommended at this time: combinations based on<br>irinotecan, cisplatin, doxorubicin |
| **PIPAC** | Oxaliplatin<br>Cisplatin/doxorubicin |
| **IP/EPIC** | 5-fluorouracil (FUDR) used in some centers |

\* Use of regional perfusion chemotherapy is extremely institution- and setting-specific, and there is neither adequate literature nor strong consensus as to the most effective regimen or mode of administration. This table is included for reference into current practices at time of writing.

## BASIC PRINCIPLES OF PATHOLOGY OF APPENDICEAL TUMORS

An overview of challenging issues in appendix tumor pathology will be described in the second part of these guidelines alongside the guideline for peritoneal disease. Key points are summarized here.

Critical points of differentiation that apply generally to pathologic evaluation of appendiceal tumors are those that are most likely to be misclassified, and those that lead to clinically relevant management differences. Expert pathology review should generally be pursued any time patients are referred from other systems to referral centers, when there is significant discordance between primary and peritoneal findings, and when signet ring cells are identified. Clinically relevant distinctions that should be closely assessed include the presence of an invasive component, which differentiates between LAMN and well-differentiated adenocarcinoma in lower grade lesions, and HAMN and adenocarcinoma in higher grade lesions. The latter should be particularly closely examined because it is common for an invasive component of HAMN to be missed.^122–124^ Well-differentiated mucinous adenocarcinoma must also be cautiously designated, as it affects the recommended extent of surgical resection.

While localized disease, by definition, should not include significant gross peritoneal disease, there may be surface mucin or perforation in specimens from disease presentations initially evaluated by surgeons as localized. Correct identification of the cellularity of extra-appendiceal mucin is critical to determine whether disease should be treated according to the peritoneal management pathway.

## APPENDICEAL MUCINOUS NEOPLASMS

### Consensus Updates

One major change from the 2018 guideline (Figure 2) is more definitive recommendations for surveillance vs cytoreduction with or without IPCT in disease that is otherwise localized but with limited regional spread. More specific recommendations address positive margins and perforation, with an emphasis on the most minimally invasive treatment possible to achieve negative margins.

**Table 4.** Delphi 1 agreement tables (% agreement includes agree and strongly agree)

|  | Strongly agree | Agree | Neither agree nor disagree | Disagree | Strongly disagree | Total | % Agree |
| --- | --- | --- | --- | --- | --- | --- | --- |
| **Block 1** | 91 | 38 | 5 | 4 | 0 | 138 | 93% |
| **Block 2** | 104 | 30 | 4 | 0 | 0 | 138 | 97% |
| **Block 3** | 92 | 37 | 7 | 2 | 0 | 138 | 93% |
| **Block 4** | 93 | 37 | 7 | 1 | 0 | 138 | 94% |
| **Block 5** | 90 | 37 | 4 | 7 | 0 | 138 | 92% |
| **Block 6** | 94 | 40 | 4 | 0 | 0 | 138 | 97% |
| **Block 7** | 93 | 39 | 4 | 2 | 0 | 138 | 96% |
| **Block 8** | 93 | 38 | 5 | 2 | 0 | 138 | 95% |
| **Block 9** | 94 | 38 | 4 | 1 | 1 | 138 | 96% |
| **Block 10** | 99 | 33 | 5 | 0 | 1 | 138 | 96% |
| **Block 11** | 83 | 42 | 9 | 4 | 0 | 138 | 91% |

**Table 5.**
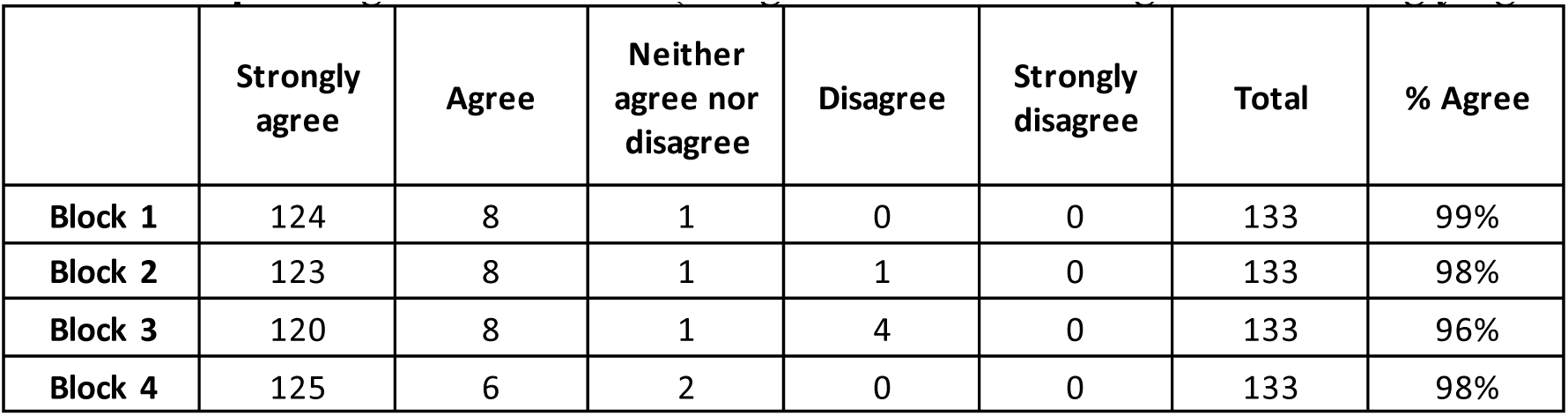
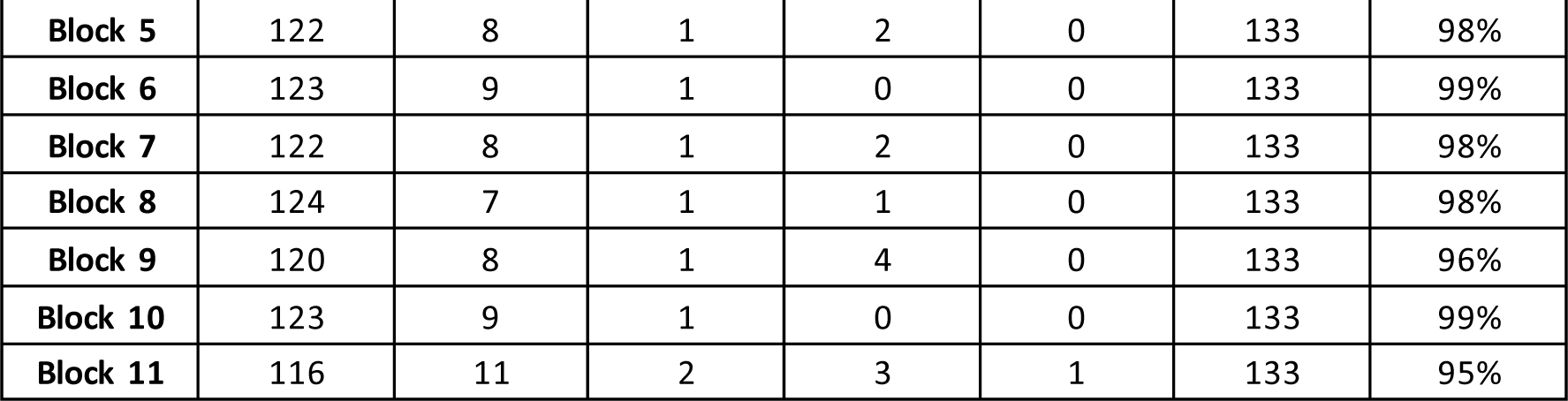
Delphi 2 agreement tables (% agreement includes agree and strongly agree)

**Figure 2.**
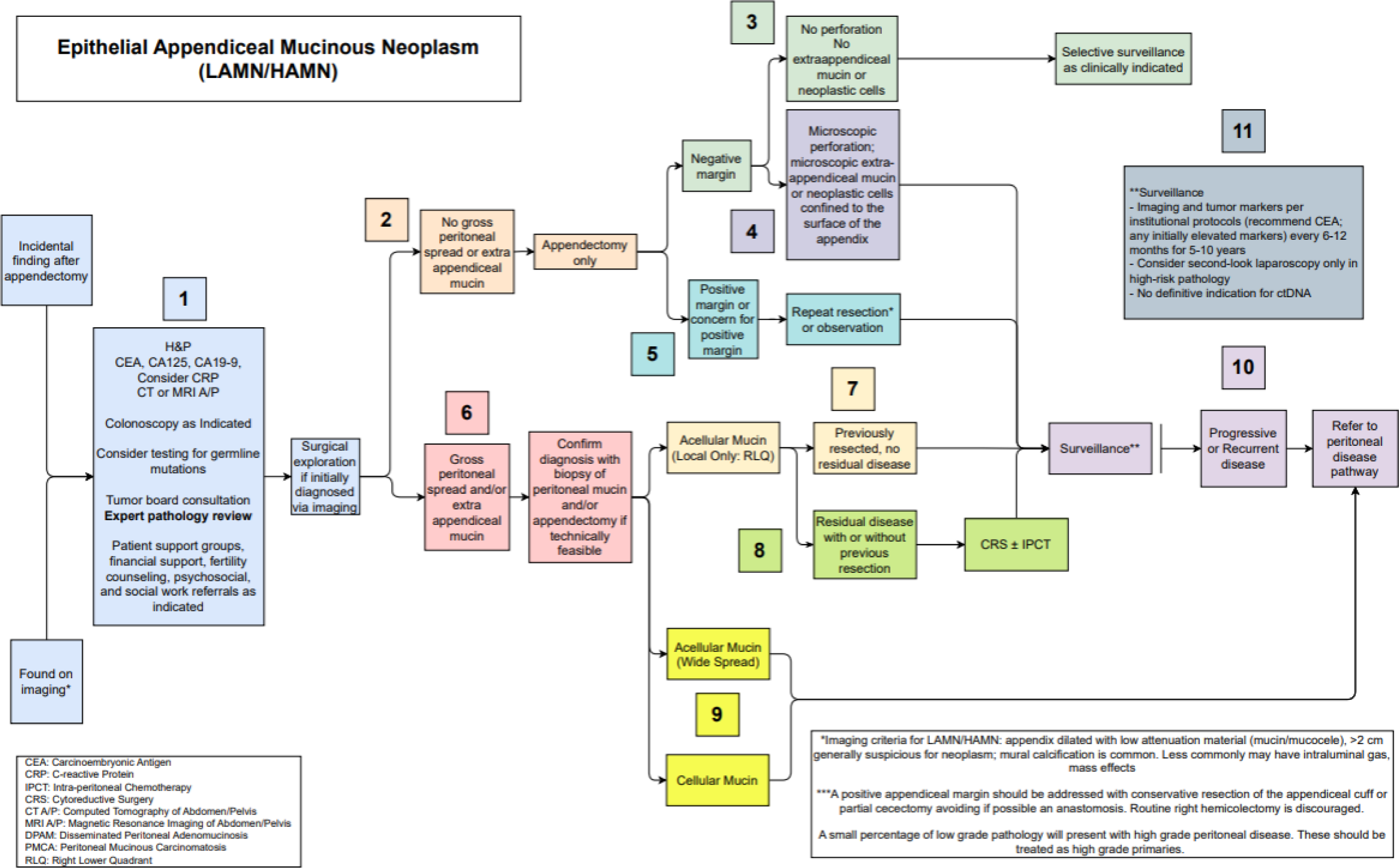
Epithelial Appendiceal Mucinous Neoplasm Pathway.

#### Block 1

When first detected on imaging, or as a pathological finding during or after appendectomy, initial workup of a suspected of AMN should include a detailed history and physical, tumor markers including CEA, CA125, CA19-9, and CRP, and abdominopelvic cross-sectional imaging if not already performed. Serum markers are useful for prognostication, monitoring treatment response, and identifying recurrence. Imaging is additionally useful for evaluating peritoneal and other distant disease sites and surgical planning.^125–128^ Imaging findings that may be seen in appendiceal neoplasms include focal distal appendiceal dilatation, size over 2 cm, curvilinear calcifications, wall irregularity, and absence of periappendiceal fat stranding; calcifications are specific but not sensitive.^129,130^

Colonoscopy should be performed to rule out synchronous lesions that might affect surgical planning, which occur in 14-42% of this population.^131,132^ Somatic and tumor genetic profiling may be considered but minimal evidence exists for AMN.

Patients with AMNs should be discussed at multidisciplinary tumor board; while many AMNs can be treated with resection alone, imaging and treatment plan review can help prepare the care team for unexpected contingencies. Tissue samples should be reviewed by an expert pathologist. Patients should also be evaluated for additional support needs, which may include referral to patient support groups, social work consultation, financial support resources, psychosocial support resources, and fertility counseling.

Where AMNs are diagnosed by any non-surgical means (typically imaging), the next step should be surgical exploration by least invasive safe approach. In most cases this will be diagnostic laparoscopy, but the surgeon’s best judgement must be employed. If lesions suspicious for peritoneal disease are identified, biopsies should be taken.

**% Agreement**: First round 93%, second round 99%

#### Block 2

If no gross peritoneal spread of disease or macroscopic extra-appendiceal mucin is noted on surgical exploration, appendectomy alone should be performed to a negative _margin. 133–135_

**% Agreement**: First round 97%, second round 98%

#### Block 3

If final surgical margins are negative, attention must be turned to the presence or absence of perforation and extra-appendiceal mucin or neoplastic cells. If all of the above are absent, surveillance can be employed selectively. In many cases surveillance will not be necessary; however, the risk of recurrence is never zero, as it is possible for an AMN to perforate and then re-seal, leading to a theoretical increased risk of peritoneal progression or recurrence.^134–136^

**% Agreement**: First round 93%, second round 96%

#### Block 4

If final surgical margins are negative but microscopic perforation is noted, or there is microscopic extra-appendiceal mucin or neoplastic cells confined to the surface of the appendix, surveillance is indicated as described in block 11. ^134–136^ Microscopic extra-appendiceal mucin and neoplastic cells confined to the surface of the appendiceal specimen alone still constitute a negative margin.

**% Agreement**: First round 94%, second round 98%

#### Block 5

If final surgical margins are positive, with viable neoplastic epithelial cells at the margin (not acellular mucin alone) or there is concern for the same, repeat resection should be performed to a negative margin, although data suggests in some series that even gross resection may be adequate.^134–136^ Historically, ileocecectomy or cecectomy have been performed, but the consensus recommendation is to perform the most conservative resection possible, such as cuff resection. Anastomosis should be avoided if possible. Then surveillance must be performed regularly. Observation may be considered for those patients at high risk for surgical morbidity, in whom there may be less benefit from oncologic resection.

**% Agreement**: First round 92%, second round 98%

#### Block 6

If, on index surgical exploration, gross peritoneal spread or extra-appendiceal mucin is noted, a definitive diagnosis must be confirmed. Biopsy of the sites of peritoneal spread and appendectomy should be performed if technically feasible, such that pathologic review can clearly confirm diagnosis and disease grade to guide therapy.

**% Agreement**: First round 97%, second round 99%

#### Block 7

If extra-appendiceal disease is limited to localized acellular mucin only by direct visualization, and all disease is completely resected (the equivalent of a complete/adequate cytoreduction), no further surgical management is indicated. Rate of recurrence is as low as 4%.^137^ The definition of localized acellular mucin is ultimately dependent upon intraoperative surgeon judgement, but expert consensus recommends defining this as disease limited to the meso-appendiceal fold and peri-appendiceal recesses. Regular surveillance is indicated and should follow the recommendations in block 11.^138^

**% Agreement**: First round 96%, second round 98%

#### Block 8

If extra-appendiceal disease is limited to acellular mucin in the right lower quadrant, but residual disease is left at the time of initial exploration with or without an attempt at resection (such as in those patients referred from outside institutions or with otherwise previous incomplete cytoreduction), evaluation should be initiated for cytoreduction with or without intraperitoneal chemotherapy; given the limited data on recurrence in this subpopulation, this is primarily an expert consensus-based recommendation.

**% Agreement**: First round 95%, second round 98%

#### Block 9

If surgical exploration reveals extra-appendiceal acellular mucin that is more widely disseminated than the peri-appendiceal region, or cellular mucin, refer to the peritoneal disease pathway, as a more comprehensive approach focused on regionally advanced disease must be pursued. Recurrence estimates for localized cellular mucin (any grade) range widely from 33-75%, comparable to disseminated disease, justifying a more aggressive approach.^137,139^

**% Agreement**: First round 96%, second round 96%

#### Block 10

If there is evidence of recurrent or progressive disease during surveillance, this would be, by definition, peritoneal disease, and care should progress to the peritoneal pathway.

**% Agreement**: First round 96%, second round 99%

#### Block 11

When indicated, surveillance should include regular interval history and physical as well as imaging and tumor markers. Either CT or MRI are acceptable; modality should be chosen for consistency and expertise in institutional practice as no clear evidence identifies a the superior exam. Tumor markers should include CEA and any other markers that are noted to be elevated at initial evaluation, or at any point in treatment. No studies provide strong evidence for duration and frequency, but a single retrospective study from the US HIPEC collaborative demonstrated imaging surveillance every 6 to 12 months to be non-inferior to more frequent schedules.^140^ Recurrence is most common within approximately the first three years postoperatively, and plateaus at approximately six years.^141,142^ Consensus therefore recommends surveillance every 6 to 12 months for 5 to 10 years; higher-grade lesions and any degree of peritoneal involvement are indications for more intense surveillance.

As cross-sectional imaging is not sensitive for early peritoneal disease, high-risk pathologic features may merit second-look laparoscopy in select cases, but this should not be pursued for the majority of patients with AMNs.^140^ There is no definitive indication for ctDNA surveillance in AMNs.

**% Agreement**: First round 91%, second round 95%

## APPENDICEAL ADENOCARCINOMA

### Consensus Results and Updates

This pathway (Figure 3) summarizes recommendations for both mucinous and non-mucinous tumors, inclusive of goblet cell but exclusive of neuroendocrine tumors. In addition to reorganization of peritoneal disease, other changes include updated criteria for systemic chemotherapy, a more comprehensive initial workup, and cohesive surveillance recommendations.

**Table 6.** Delphi 1 agreement tables (% agreement includes agree and strongly agree)

|  | Strongly agree | Agree | Neither agree nor disagree | Disagree | Strongly disagree | Total | % Agree |
| --- | --- | --- | --- | --- | --- | --- | --- |
| **Block 1** | 98 | 34 | 2 | 3 | 1 | 138 | 96% |
| **Block 2** | 102 | 28 | 5 | 2 | 1 | 138 | 94% |
| **Block 3** | 98 | 28 | 6 | 6 | 0 | 138 | 91% |
| **Block 4** | 102 | 31 | 5 | 0 | 0 | 138 | 96% |
| **Block 5** | 100 | 34 | 4 | 0 | 0 | 138 | 97% |
| **Block 6** | 99 | 33 | 5 | 1 | 0 | 138 | 96% |
| **Block 7** | 89 | 35 | 7 | 7 | 0 | 138 | 90% |

**Table 7.** Delphi 2 agreement tables (% agreement includes agree and strongly agree)

|  | Strongly agree | Agree | Neither agree nor disagree | Disagree | Strongly disagree | Total | % Agree |
| --- | --- | --- | --- | --- | --- | --- | --- |
| **Block 1** | 124 | 8 | 0 | 1 | 0 | 133 | 99% |
| **Block 2** | 124 | 5 | 1 | 3 | 0 | 133 | 97% |
| **Block 3** | 123 | 8 | 2 | 0 | 0 | 133 | 98% |
| **Block 4** | 123 | 8 | 1 | 1 | 0 | 133 | 98% |
| **Block 5** | 127 | 5 | 1 | 0 | 0 | 133 | 99% |
| **Block 6** | 122 | 10 | 1 | 0 | 0 | 133 | 99% |
| **Block 7** | 116 | 11 | 3 | 2 | 1 | 133 | 95% |

**Figure 3.**
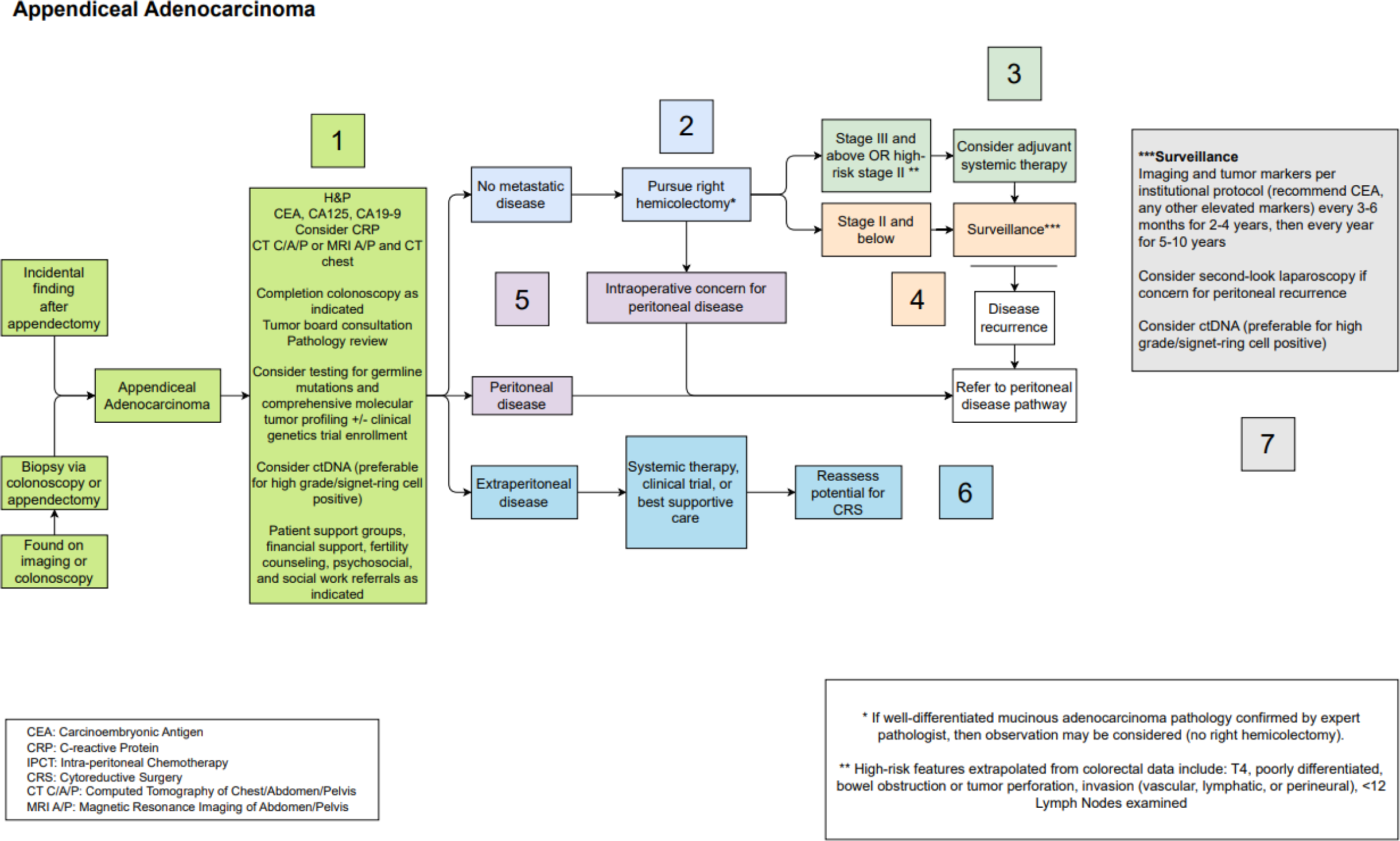
Appendiceal Adenocarcinoma Pathway.

#### Block 1

As with AMN, appendiceal adenocarcinoma may be detected on diagnostic imaging or incidentally following appendectomy. Initial evaluation and management should mirror that of the AMN pathway. ^131,132^ As discussed above, germline testing may be considered in conjunction with family cancer history for research purposes and assessment of hereditary cancer risk.^89,143–145^ Comprehensive tumor profiling should be considered to identify potential molecular targets. ^125–128,143,144^ Of note, ctDNA testing may be considered particularly for patients with high-grade or signet ring cell pathology as it is useful for prognostication, although evidence is limited in appendix cancer compared to metastatic colorectal cancer.^146^

**% Agreement**: First round 96%, second round 99%

#### Block 2

Right hemicolectomy (RHC) with oncologic lymphadenectomy should be pursued for most cases of appendiceal adenocarcinoma in suitable surgical candidates. Currently this is interpreted as the 12-node yield required in colon cancers. Observational data shows survival benefit with at least 10 nodes.^147^ Although stage migration and limitations of current research may contribute to the observed benefit of RHC, it has been associated with survival benefit in most mucinous adenocarcinomas with a stage greater than 1, and any non-mucinous adenocarcinoma.^148,149^

The exception to this is well-differentiated mucinous adenocarcinoma that is completely confined to the appendix with negative margins and no concern for more distant disease. The rate of lymph node positivity has been shown to be low in well- and some moderately-differentiated mucinous lesions, decreasing the survival benefit of RHC.^72,133,150^

**% Agreement**: First round 94%, second round 97%

#### Block 3

Patients with stage III appendiceal adenocarcinoma (spread to at least one regional lymph node) or stage II appendiceal adenocarcinoma with any high-risk features should be considered for adjuvant systemic chemotherapy following surgical resection.^52,54,60,72,72,73,73–78^ High-risk features are summarized in the systemic chemotherapy section above.^52,54,71–77,147^ Adjuvant chemotherapy regimens, described under the systemic chemotherapy section, typically last 3-6 months, depending on patient toleration, with a goal of 6 months of therapy. ^26,32,57^ Patients should be subsequently surveilled, as described in Block 7.

**% Agreement**: First round 91%, second round 98%

#### Block 4

Patients with stage I and II appendiceal adenocarcinoma without high-risk features as defined above, should be surveilled following surgical resection, as described in block 7, as there is insufficient evidence to suggest that systemic chemotherapy is beneficial in low risk lesions after complete resection.^71^

**% Agreement**: First round 96%, second round 98%

#### Block 5

If recurrent disease is detected on initial diagnostic workup or during surgical resection, management should follow the pathway described for appendiceal tumors with peritoneal disease, which will be presented separately and will address both peritoneal and extra-peritoneal disease.

**% Agreement**: First round 97%, second round 99%

#### Block 6

Although not an absolute contraindication to resection in oligometastatic disease, appendiceal adenocarcinoma with extraperitoneal spread at diagnosis is a poor prognostic indicator, and patients presenting in this setting are unlikely to be candidates for definitive surgical resection.^151^ Through joint decision-making, clinicians and patients may consider systemic chemotherapy, clinical trials, or best supportive care alone. Multidisciplinary oncologic care including considering palliative consultation is recommended. Surgical intervention may be appropriate for symptom control. Depending on response to intervention, patients may be re-evaluated for debulking or more definitive cytoreductive surgery.

**% Agreement**: First round 96%, second round 99%

#### Block 7

Imaging and clinical surveillance with the same elements as for AMN is recommended at a frequency of every 3-6 months for 2-4 years, followed by annually for 5 to 10 years. This is more frequent than recommended for AMNs, given the higher recurrence rates in this population in the first year after resection, but is similar to surveillance for higher grade colorectal disease. ^140–142^ As with AMN, cross-sectional imaging is not sensitive for early peritoneal disease, thus second-look laparoscopy may be considered where there is concern for peritoneal recurrence.^140^ Again, interval testing for circulating tumor DNA levels should also be considered, particularly for patients with high-grade or signet-ring positive pathology.^146^

**% Agreement**: First round 90%, second round 95%

## DISCUSSION

This text summarizes two of three consensus guideline pathways regarding the management of appendiceal tumors without peritoneal involvement. Consensus was achieved after two rounds of review by a multidisciplinary group across all pathway blocks.

Most evidence regarding the treatment of appendiceal malignancy remains observational at best; however, the volume of data has increased, and there is incrementally improving understanding of the role of systemic chemotherapy. One of the chief benefits of this update is unification of recommendations both across consensus group members with multiple different roles in the comprehensive cancer care field, and across a single unified pathologic grading system. Major changes to emphasize in localized disease recommendations are the new preferential recommendations for margin resection only for LAMN (avoiding segmental resections and anastomoses where possible) and clarified recommendations regarding chemotherapy.

Limitations of the consensus include the retrospective and observational nature of almost all relevant literature in appendiceal neoplasm management. The role of intraperitoneal chemotherapy remains highly controversial among consensus members, and thus no explicit recommendation is presented here. The increased diversity in expertise represented in this consensus group is a major strength.

### Comparison to other international guidelines

Both the American Society of Colorectal Surgeons (2019) and the Peritoneal Surface Oncology Group International (2021) have published their own consensus guidelines since the development of the Chicago Consensus, but both have limitations.^152,153^ The ASCRS guidelines are surgeon-focused, while PSOGI guidelines are more relevant to the European practice environment and do not expand upon certain grade-by-grade distinctions in management which have been demonstrated to be clinically relevant. The PSOGI guidelines focus on peritoneal disease but also include some guidelines relevant to localized disease as here. First and foremost, they sit within the larger ecosystem of common PSOGI terminology and rely on the PSOGI pathologic classification system.^152^ Initial evaluation guidelines are similar to this consensus, except of CRP or any genetic workup is not recommended. Surgical recommendations, including trocar placement (midline to allow for port excision), are more specific, although diagnostic laparoscopy is not as strongly recommended prior to resection; our consortium guidelines essentially require tissue diagnosis.

PSOGI presents recommendations separately for goblet-cell adenocarcinoma (GCA), instead of their inclusion in this consensus along with other non-mucinous adenocarcinoma. PSOGI suggests hemicolectomy may not be necessary in the lowest grade tumors (WHO grade 1 GCA or Tang A) confined to the appendix without high-risk features, while our group at this time recommends right hemicolectomy without exception. Conversely, PSOGI supports consideration of right hemicolectomy for HAMN even without peritoneal disease, while the PSM consortium favors resection to negative margins only. The PSOGI consensus also suggests perforation may be an indication to consider cytoreduction, but our group recommends cytoreduction only if there is demonstrable peritoneal disease of either cellular character or outside the immediate peri-appendiceal region. In terms of systemic chemotherapy, PSOGI specifically recommends a 5-FU backbone and an alkylating agent, as well as neo-angiogenesis inhibitors where resection is incomplete or not performed; indications are generally similar although no preference is given for preoperative vs postoperative timing.^152^

The ASCRS guidelines are from a focused surgical perspective with some salient differences, including that no exception to hemicolectomy is made for well-differentiated and otherwise localized adenocarcinoma. Recommendations regarding systemic chemotherapy are very limited and only extend to unlikely benefit in low-grade lesions and possible benefit in HAMN.^153^

The Peritoneal Malignancies Oncoteam of the Italian Society of Surgical Oncology recently published recommendations as well. Recommendations overlap in most areas with PSOGI, including using PSOGI terminology, considering hemicolectomy for HAMN, and pursuing cecectomy or ileocecectomy for margin involvement in LAMN instead of conservative margin resection alone. Similarly, where cellularity or dissemination of peritoneal mucin is required to consider cytoreduction in the PSM consortium guidelines, perforation alone is grounds for considering CRS/HIPEC in the SICO consensus.^154^

### Patient perspective

Advocacy groups such as PMP Pals and Appendix Cancer Pseudomyxoma Peritonei Research Foundation (ACPMP) are key resources both directly to patients and families, and indirectly by engaging with research initiatives and guiding clinical practice. Diagnosis with rare malignancies such as appendix tumors often leaves patients and their caregivers feeling abandoned and without options. Moreover, the process of treating appendiceal cancer is far from benign, with long-lasting effects on physical, sexual, and mental health for which patients and families are often not adequately prepared. Respondents identified strong community as crucial to alleviating those feelings, including close relationships with a network of oncologists, surgeons, advocacy groups, and family, and for some, integration of alternative, holistic, and palliative practitioners into routine care. The multidisciplinary nature of this consensus seeks to produce a cohesive approach that facilitates an integrated support network.

Responses from advocacy groups emphasize that quality of life and survival are paramount in deciding on treatment modalities, but that those decisions are not always obvious, especially during surveillance following surgery. One respondent described the experience as a “vast wasteland,” with patients “left to wander a five-year journey with little on the horizon.” Well-designed, accessible online resources are key roadmaps for many, while it is access to clinical trials that often provides direction to that journey by offering hope and a sense of autonomy. However, some patient and caregiver advocates report struggling to navigate this process due to constraints of geography and medical insurance. While this guideline emphasizes referral to clinical trials, equitable access to trials for all has not been achieved. Patient advocates emphasize that current research and scholarship involving appendiceal malignancies would benefit from a louder patient voice, whether it be in choice of study design, deciding on outcomes of interest, or educational initiatives and communication.

Ultimately, improving patients’ experience hinges on clarifying the treatment journey, limiting isolation, and fostering hope where possible.

### Future scope

Recommendations related to systemic chemotherapy are in need of ongoing study as outcomes remain poor, particularly in patients with high grade disease. A clear need remains for judiciously designed prospective trials to identify the optimal sequence and delivery of treatment modalities for patients with appendiceal tumors; some are in current development, particularly to investigate the neoadjuvant setting. Most randomized trial schemata are difficult to employ in this patient population, but recent work using crossover designs has shown promise.^68^

Further work is needed to explore quality of life outcomes for patients with appendix tumors, as the relative rarity of their disease leaves them with less support than individuals facing more common cancers.

## CONCLUSIONS

In conclusion, herein is reported an updated Delphi consensus of management guidelines concerning appendiceal tumors without peritoneal involvement. Importantly, this consensus group contained experts across multiple disciplines relevant to cancer care, including medical oncologists, surgical oncologists, pathologists, radiologists, palliative care specialists, and patient advocates. Surgical resection remains the primary modality of up-front definitive treatment in presentations without peritoneal involvement. Systemic chemotherapy should be considered for high-risk pathologies. Regular surveillance should be performed for all patients with appendiceal tumors, save the lowest-grade, lowest risk LAMNs after complete resection with no additional risk factors.

## Data Availability

The datasets used and/or analysed during the current study are available from the corresponding author on reasonable request.

## Notes

### Competing Interest Statement

The authors have declared no competing interest.

### Clinical Protocols

https://www.crd.york.ac.uk/prospero/display_record.php?RecordID=463216

### Funding Statement

This study did not receive any funding.

