## Supplement Tables 1-3 for "Consensus guideline for the management of patients with appendiceal tumors Part 1: Appendiceal tumors without peritoneal involvement"

**SUPPLEMENTARY MATERIAL**

**Supplemental Table 1. Search Strategy for Key Question 1**

PubMed; restrictions: any date - 2023/06/15, performed on humans

| Search line | Search term |
|---|---|
| 1 | Appendi*[tw] AND (cancer*[tw] OR adenocarcinoma*[tw] OR carcinoma*[tw] OR neoplasm*[tw]) OR "goblet cell"[tw] OR (Appendiceal neoplasms[MeSH Major Topic]) OR pseudomyxoma[tw] OR "pseudomyxoma peritonei"[MeSH Major Topic] |
| 2 | ("surgery peritoneal"[tiab:~5]) OR ("surgery peritoneum"[tiab:~5]) OR ("resection peritoneal"[tiab:~5]) OR ("resection peritoneum"[tiab:~5]) OR (Cytoreduc*[tw] OR CRS[tw]) |
| 3 | (Chemotherapy OR Immunotherapy[tw] OR Targeted therapy[tw] OR Systemic therapy[tw] OR Drug Therapy[MeSH Major Topic] OR Antineoplastic agents[MeSH Major Topic] OR Drug therapy, Combination[MeSH Major Topic] OR chemotherapy, adjuvant[MeSH Major Topic] OR neoadjuvant therapy[MeSH Major Topic]) |
| 4 | #1 AND #2 AND #3 |

**Supplemental Table 2. Inclusion Criteria and Review Materials**

| REASON | CRITERIA | EXCLUSIONS |
|---|---|---|
| **Language/availability** | Available in English | Not able to obtain abstract in English |
| **Study design** | Original research<br>Cohort or case-control study, or single-arm or randomized interventional trial | Case series or report<br>Population overlaps with another study/population and cannot be clearly delineated |
| **Population** | Individuals with appendix tumors with peritoneal involvement, other than low-grade (well-differentiated), undergoing first-time treatment for PSM including cytoreductive surgery | Population does not have peritoneal disease, appendiceal origin, or moderately-/poorly-differentiated histology and/or is not undergoing cytoreduction<br>Chemotherapy administered without plan for cytoreduction |
| **Intervention** | Any regimen and any timing of systemic chemotherapy, other than a single dose with IPCT intraoperatively | No description or sub-analysis of receipt of chemotherapy<br>Other interventions are present that would confound analysis (other surgeries, cancers) |
| **Comparator** | No comparator required for raw survival time<br>Can compare to alternative regimens of chemotherapy or none (CRS alone)<br>If outcomes restricted to comparative metrics, must be compared to similar CRS patients with an eligible chemotherapy regimen or none | Comparative metrics only, with ineligible comparator group |
| **Outcome** | Outcomes of interest: survival, recurrence, progression, adverse/treatment effects | No description or sub-analysis of outcomes by treatment group |

### Supplemental Table 3. Quality Assessment of Included Studies

| Supplemental Table 3. Quality assessment of studies assessing systemic chemotherapy regimens and timing relative to cytoreduction in peritoneal appendiceal malignancy | | | | |
|---|---|---|---|---|
| **Author** | **Selection** | **Comparability** | **Outcomes** | **Overall** |
| Barrak 2021[1] | **** | - | *** | ******* |
| Sugarbaker 2021[2,3] | **** | - | ** | ****** |
| Sugarbaker 2010[3] | *** | - | *** | ****** |
| Sugarbaker 2022[4] | **** | - | *** | ******* |
| Bijelic 2012[5] | **** | - | * | ***** |
| Ihemelandu 2016[6] | **** | ** | * | ******* |
| Sugarbaker 2023[7] | **** | ** | *** | ********* |
| Mangieri 2022[8] | **** | ** | * | ******* |
| Votanopoulos 2015[9] | **** | - | ** | ****** |
| Blackham 2014[10] | **** | * | *** | ******** |
| Cummins 2016[11] | **** | ** | ** | ******** |
| Munoz-Zuluaga 2019[12] | **** | ** | *** | ********* |
| Baron 2023[13] | **** | ** | *** | ******** |
| Milovanov 2015[14] | **** | ** | *** | ********* |
| Munoz-Zuluaga 2019[15] | **** | ** | ** | ******** |
| Morgan 2023[16] | **** | ** | * | ******* |
| Kolla 2020[17] | **** | * | * | ****** |
| Chen 2020[18] | **** | ** | * | ******* |
| Flood 2023[19] | **** | ** | * | ******* |
| Turner 2013[20] | **** | - | *** | ******* |
| Hanna 2023[21] | **** | ** | ** | ******** |
| Spiliotis 2017[22] | **** | * | * | ****** |
| Benhaim 2019[23] | **** | ** | ** | ******** |
| Mercier 2019[24] | **** | - | *** | ******* |
| Masckauchan 2019[25] | **** | ** | ** | ******** |
| Acs 2023[26] | **** | ** | ** | ******** |
| Kusamura 2021[27] | **** | ** | ** | ******** |
| Baratti 2008[28] | **** | ** | *** | ********* |
| Schomas 2009[29] | **** | ** | ** | ******** |
| Arjona-Sanchez 2013[30] | **** | * | *** | ******** |
| Ung 2014[31] | **** | ** | * | ******** |
| Baumgartner 2015[32] | **** | ** | * | ******* |
| Grotz 2017[33] | **** | ** | ** | ******** |
| Pallas 2017[34] | **** | ** | * | ******* |
